# Lesion Volume as a Predictor for Return to Work After Endovascular Therapy – a Four-year Prospective Cohort Study

**DOI:** 10.1101/2024.04.16.24305935

**Authors:** Gisle Berg Helland, Mona Kristiansen Beyer, Brian Anthony B. Enriquez, Hege Ihle-Hansen, Håkon Ihle-Hansen, Stein Andersson, Esten Høyland Leonardsen, Helle Stangeland, Bettina Ùjhelyi, Guri Hagberg, Hanne Flinstad Harbo, Anne Hege Aamodt, Einar August Høgestøl

## Abstract

**Background:** There are no known objective biomarkers for predicting Return to Work (RTW) in ischemic stroke survivors. This study aims to explore the predictive utility and define a cut-off value of lesion volume on RTW after endovascular treatment (EVT).

**Methods:** We included patients <65 years undergoing EVT at Oslo University Hospital (OUS) between January 2017 and May 2019. Employment status was obtained at both baseline and a four-year follow-up. Stroke lesion volumes were segmented using magnetic resonance imaging (MRI) scans 24 hours post EVT. Logistic regression models were conducted to assess the impact of lesion volume on RTW-status at follow-up, adjusted for patients’ characteristics, stroke related factors and treatment. We calculated the Receiver Operating Characteristic curve to determine the optimal lesion volume cut-off. Machine learning (ML) regression models were employed to assess the predictive abilities of baseline clinical and imaging variables for RTW.

**Results:** Of the 109 individuals treated, 81 (74%) were employed at baseline. Among these, 60 completed four-year follow-up with MRI available for stroke lesion segmentation and were included in the analyses. Mean age at stroke onset was 51.8 years (range 23.5–64.9), 50% were female. Median lesion volume was 18 ml (IQR 45-7). After four years, 34 (57%) had successfully RTW. The odds for not RTW increased by 5% for every 1 ml increase in lesion volume (adjusted odds ratio [aOR], 95% CI, 1.02–1.11], P=0.02). A lesion volume cut-off value of 29 cm^3^ yielded a sensitivity of 0.91 and specificity of 0.65 for predicting RTW. Notably, the most influential feature in the ML model for predicting RTW was lesion volume.

**Conclusion:** Lesion volume was the most robust predictor of RTW four years after EVT. Our findings suggest that a cut-off of 29 cm^3^ is suitable to distinguish between those with high and low chance of RTW.

## Introduction

Return to work (RTW) is associated with enhanced well-being and improved quality of life for stroke-survivors.^1^ Stroke imposes a substantial burden on society, and work-related issues account for a considerable share of those costs.^2,3^ In heterogenous stroke populations, the reported rates of RTW in working-age adults vary, ranging from 7% to 81%.^4,5^ Studies focusing on patients receiving endovascular therapy (EVT) report that approximately one out of three achieve RTW within the first year after discharge.^6,7^

Several factors have been identified as determinants for successful RTW, with male sex and younger age being the most consistent protective factors.^8,9^ Time since the acute incident is also important, as there is a growing number of patients achieving RTW in the years following a stroke.^8^ Other potential prognostic factors for RTW in patients undergoing EVT include the addition of intravenous thrombolysis (IVT), successful recanalization (Thrombolysis in Cerebral Ischemia (TICI) 2b – 3), medical complications and length of hospital stay.^7^

The modified Rankin Scale (mRS) and the National Institutes of Health Stroke Scale (NIHSS) are key measures of functional post-stroke outcomes and stroke severity, and studies indicate that lower mRS and NIHSS scores during the index stay increases the likelihood of RTW.^6,8,10^ None of the factors listed above, individually, are sufficient predictors of RTW.^11,12^ In recent years, lesion volume has emerged as a potential biomarker for functional and cognitive outcome,^13–15^ but its capacity to predict RTW has not been investigated.

We aimed to explore the predictive value of lesion volume for RTW in patients undergoing EVT and establish a lesion volume cut-off to guide prognostic expectations. Secondary, we aimed to assess the predictive capabilities and influence of clinical and imaging variables on employment status four years post-EVT, employing an interpretable machine learning model.

## Methods

### Study sample and design

This study utilizes data from "The Oslo Acute Reperfusion Stroke Study" (OSCAR, study-ID: NCT06220981), a prospective cohort study of consecutive acute ischemic stroke patients treated with EVT at Oslo University Hospital (OUH). The OUH is a comprehensive stroke center receiving EVT candidates from local stroke units. In this sub-study, all patients 65 years and younger who underwent EVT between January 1, 2017, and May 9, 2019, were included. Follow-up was performed from September 2021 to June 2022. Baseline data were collected from the OSCAR study and medical records. We adhered to the STROBE guidelines when reporting our findings.^16^ The first author had access to all data and assumes responsibility for its integrity and accuracy of the data analysis.

### Imaging acquisition and imaging variables

Magnetic Resonance Imaging (MRI) scans were acquired 24 hours post EVT, at four different Siemens scanners at OUH (Table S1). Lesions were identified as areas with diffusion restriction on Diffusion Weighted Images (DWI), indicative of irreversible brain infarction.^17^ In two patients without 24-hour imaging, initial admission DWIs were used. Lesion segmentation was manually executed using the ’Insight Segmentation and Registration Toolkit-Snap’ (IKT-Snap) software (v. 4.0.0) (Figure 1).^18^ Segmentations were performed by trained researcher G.B.H and overseen by experienced neuroradiologist M.K.B, who was blinded to the outcome and clinical data.

**Figure 1.**
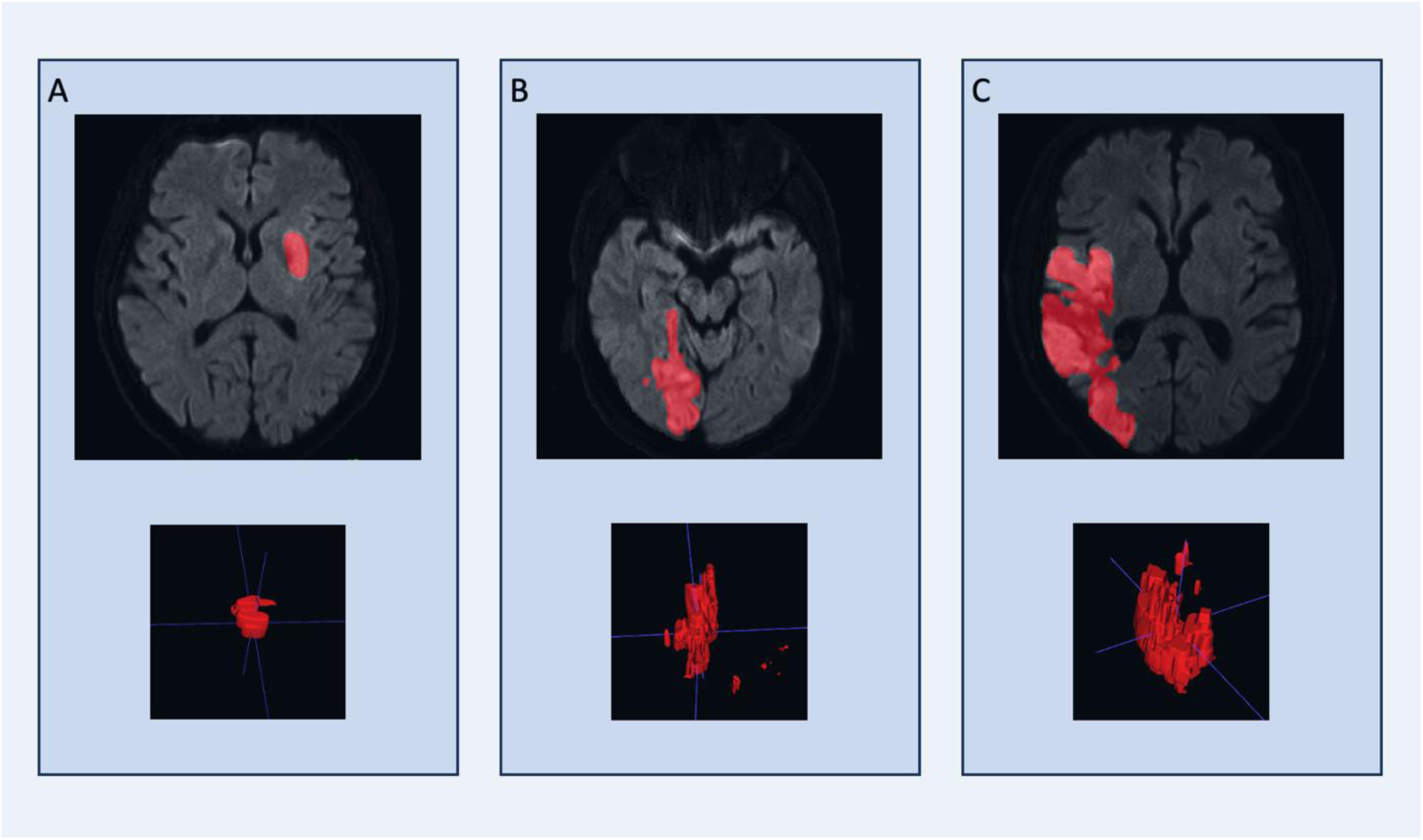
Examples of diffusion weighted images together with its corresponding lesion segmentation and categorization. **A)** Left Middle Cerebral Artery infarct, small lesion (volume 6.1 cm^3^). **B)** Posterior Cerebral Artery infarct, medium lesion (volume 22.0 cm^3^). **C)** Right Middle Cerebral Artery infarct, large lesion (volume 94.7 cm^3^).

Lesion location is grouped to left or right anterior- or posterior circulation, as described on the diagnostic CT angiography acquired at admission. Reperfusion grade was assessed using the revised modified Treatment In Cerebral Ischemia (mTICI) scoring system (score 0-3) by the interventional neuroradiologist during the final Digital Subtraction Angiography procedure.^19,20^ Missing reperfusion scores were retrospectively completed by an interventional neuroradiologist blinded for clinical data including outcomes. For analysis, mTICI scores were dichotomized, with 3 defined as an “excellent“ outcome.^21^

### Follow-up

All patients included at baseline were invited to a four-year follow-up examination. Those unable to attend in person were interviewed by telephone to collect basic demographic and clinical information. The patients were provided two self-report questionnaires related to their mental health and quality of life, including the Hospital Anxiety and Depression Scale (HADS) and the EuroQol-5Dimensions-5Levels (EQ-5D-5L).^22,23^ Fatigue was assessed using a nine-point fatigue severity scale (FSS).^24^ Information regarding adherence to work was obtained and “RTW” was defined as part or full-time paid work, or being enlisted as a student. Participants who stated that they retired early due to stroke impairments, were considered as Not Returned to Work (NRTW). Statistical comparisons between those who RTW, did NRTW and “Retired” can be found in Table S2. Information concerning education level was collected retrospectively at follow-up. For the analyses, ’primary school’ and ’high school’ were categorized into ’high school’, and ’college’ and ’university’ into ’higher education’.

### Statistical analysis

Traditional statistical analyses were performed using the R (v. 4.3.2) statistical software.^25^ Baseline characteristics are presented as number (%), medians and interquartile ranges (IQR), means and standard deviations (SD), as well as ranges, as appropriate. We conducted two-sided t-tests to analyze continuous variables that followed a normal distribution, while the Mann-Whitney U test was used for data with non-normal distribution. For categorical variables, Chi-square test and Fischer’s Exact test was used based on observed frequencies in the contingency tables.

To explore the association between lesion volume and RTW, we performed a logistic regression analysis with lesion volume as a continuous variable. Results were reported for both univariable and multivariable models, with odds ratios (OR) and 95% confidence intervals (CI). Variables selected for the univariable logistic regression analysis were based on clinical expertise and insights from previous studies. Variables included sex, age, education, affected vascular territory, NIHSS on admission and at discharge, mTICI score and whether IVT was administered or not.^8^ In the multivariable logistic regression model, sex, age, and education were predefined variables. Other variables were incorporated if they had a p-value below 0.2 in univariable analyses and variation inflation factor among variables was below 5.^26^ An interaction analysis between NIHSS and lesion volume was performed. To further validate our findings, we conducted the aforementioned logistic regressions, normalizing lesion volume to the estimated total intracranial volume, as determined by FreeSurfer, version 7.3.2.^27^ T1 weighted images taken 24 hours post-EVT were used for this analysis.

To establish a lesion volume cut-off, we made a Receiver Operating Characteristic (ROC) curve and determined the lesion volume to optimally balance sensitivity and specificity using Youden’s index.

To enhance the interpretability further, we performed a univariable logistic regression analysis, categorizing lesion volume and NIHSS scores at discharge. Lesion volumes were classified into small, medium and large (<15 cm^3^, 15-70 cm^3^ and >70 cm^3^), based on previously published cut-offs from the ESCAPE-NA1 trial, which looked at stroke lesion volume and functional outcome (mRS).^14^ NIHSS scores were categorized as minor (0-4), moderate (5-15) and severe (16-42), in line with international guidelines.^28^ Categorization of admission NIHSS scores was not performed due to the minimal number of patients falling into the reference category.

### Machine learning model

To explore the predictive properties of a larger set of baseline data using a data-driven approach, a machine learning model was developed using python with building blocks from scikit-learn.^29^ A dataset was assembled using domain expertise, comprising 28 baseline features (Table S3). While inspecting the dataset we identified some missing values (35/1740 = 2%), which were imputed with the mean of their respective features.

To establish the model, we performed a 10-fold cross-validation stratified for outcome and sex to increase the robustness of the results. Evaluation metrics included ROC and area under the curve (AUC), sensitivity, specificity, and accuracy. We utilized a LASSO regularization to perform automatic feature selection.^30^ Furthermore, we conducted a permutation test to assess the statistical significance of the predictive performance of our machine learning model compared to chance levels. In this test, we repeatedly shuffled the employment outcome variable while keeping the predictors intact. This process was repeated 100 times to establish a distribution of baseline performances of models fitted to noise, against which the performance of the true model was compared.^31^ Statistical significance was assessed by evaluating whether the mean AUC achieved by the true model was in the 95^th^ percentile of the AUC’s in the baseline distribution, equivalent to a significance threshold of 0.05. The ML modelling methodology and reporting of results in this study align with published recommendations.^32^

### Ethical approval

The study was carried out in accordance with the Helsinki Declaration and received approval from the regional ethics committee (Regional Committee for Medical Ethics Southeast Norway, (REC ID: 152864). Informed consent was obtained from all eligible study participants, and for those unable to provide consent, their next-of-kin were approached to seek consent on their behalf.

### Data availability

Supporting data for the findings in this study will be made available from the corresponding author upon reasonable request. To facilitate replicability, the pre-trained machine learning model is made publicly available at https://github.com/gislebh/Return_to_Work_after_Stroke.

## Results

A total of 416 patients underwent EVT, of whom 133 were below 65 years of age at stroke onset. Among those employed at baseline, 60 provided consent and were followed up after four years, as seen in the flow chart in Figure 2.

**Figure 2.**
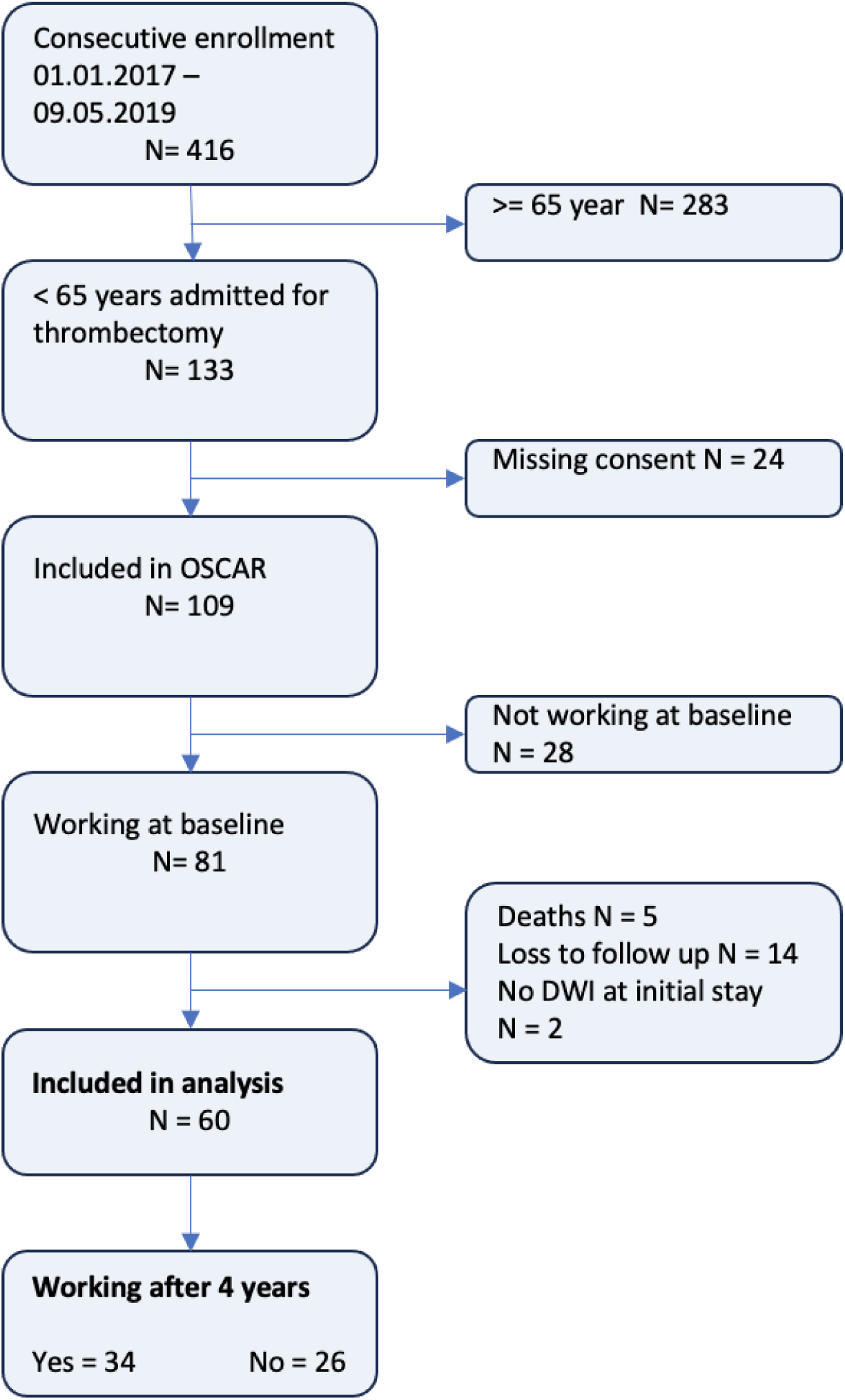
Flow chart of patient selection with an illustration of enrollment, inclusion, exclusion and loss to follow up.

Mean age at baseline was 52 years (SD 9 years, range 23.5 – 64.9). 50% were female. Median NIHSS score on admission was 12 (IQR 19-7) and decreased to 3 (IQR 7-1) upon discharge. The median lesion volume within the first 24 hours was 18 ml (IQR 45-7). The patient who RTW and had the largest lesion volume showed a volume of 79 cm^3^, whereas the patient who NRTW with the smallest lesion volume had a volume of 6.1 cm^3^.

Excellent recanalization was achieved in 21 (35%) of the participants. At the four-year follow-up, 34 (57%) participants had RTW and 26 (43%) were NRTW. Characteristics of the study population according to RTW are presented in Table 1. Figure 3 shows a box plot of lesion volume (cm^3^), including individual data, based on employment status at follow-up. Descriptive statistics of patients lost to follow-up is provided in Table S4.

**Figure 3.**
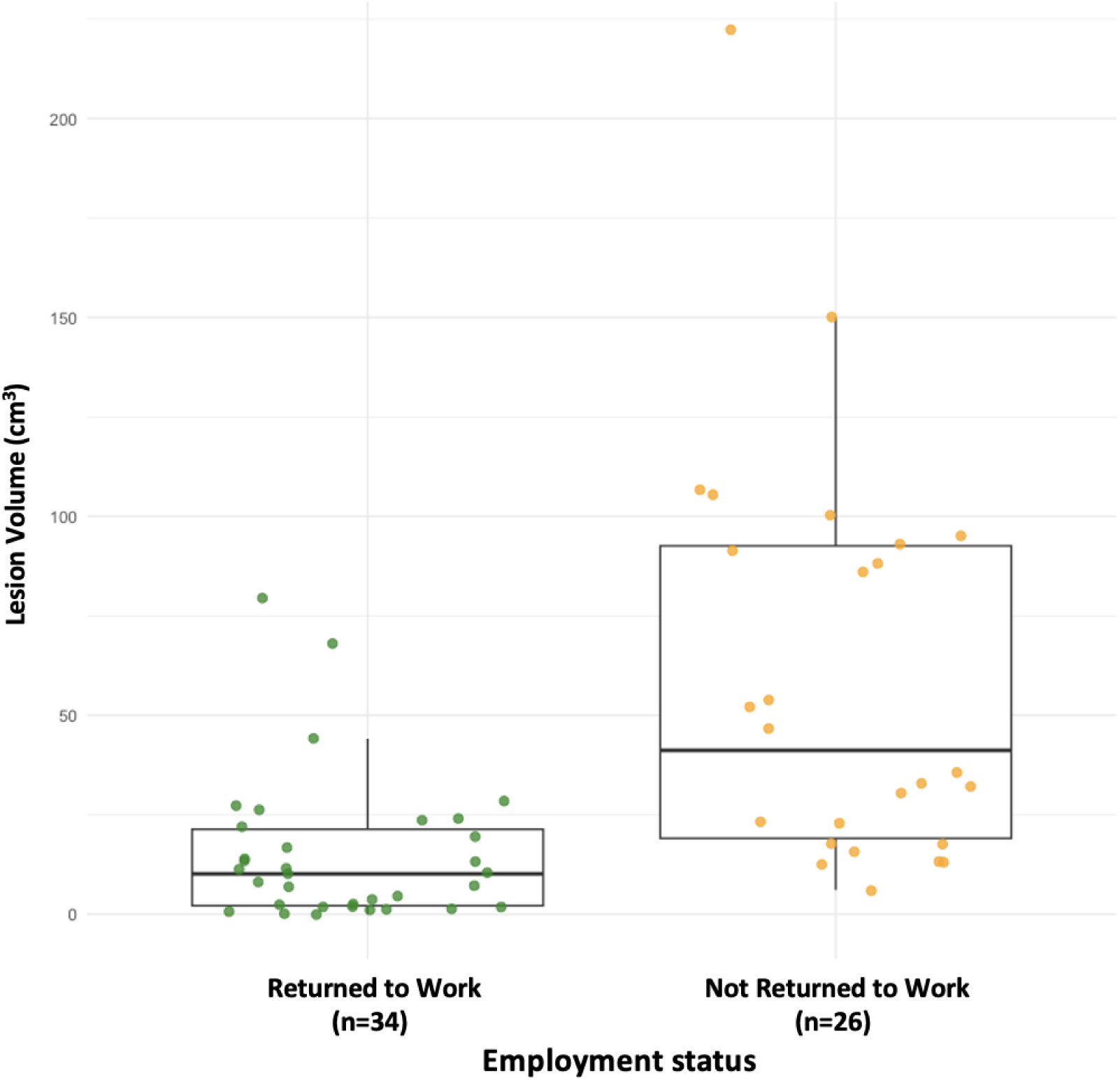
Employment status at follow up (≈ 4 years) after stroke and lesion volume at baseline, here visualized through a box plot with individual data, including median and interquartile range. Green indicate subjects who returned to work, and orange those who did not return to work.

**Table 1:**
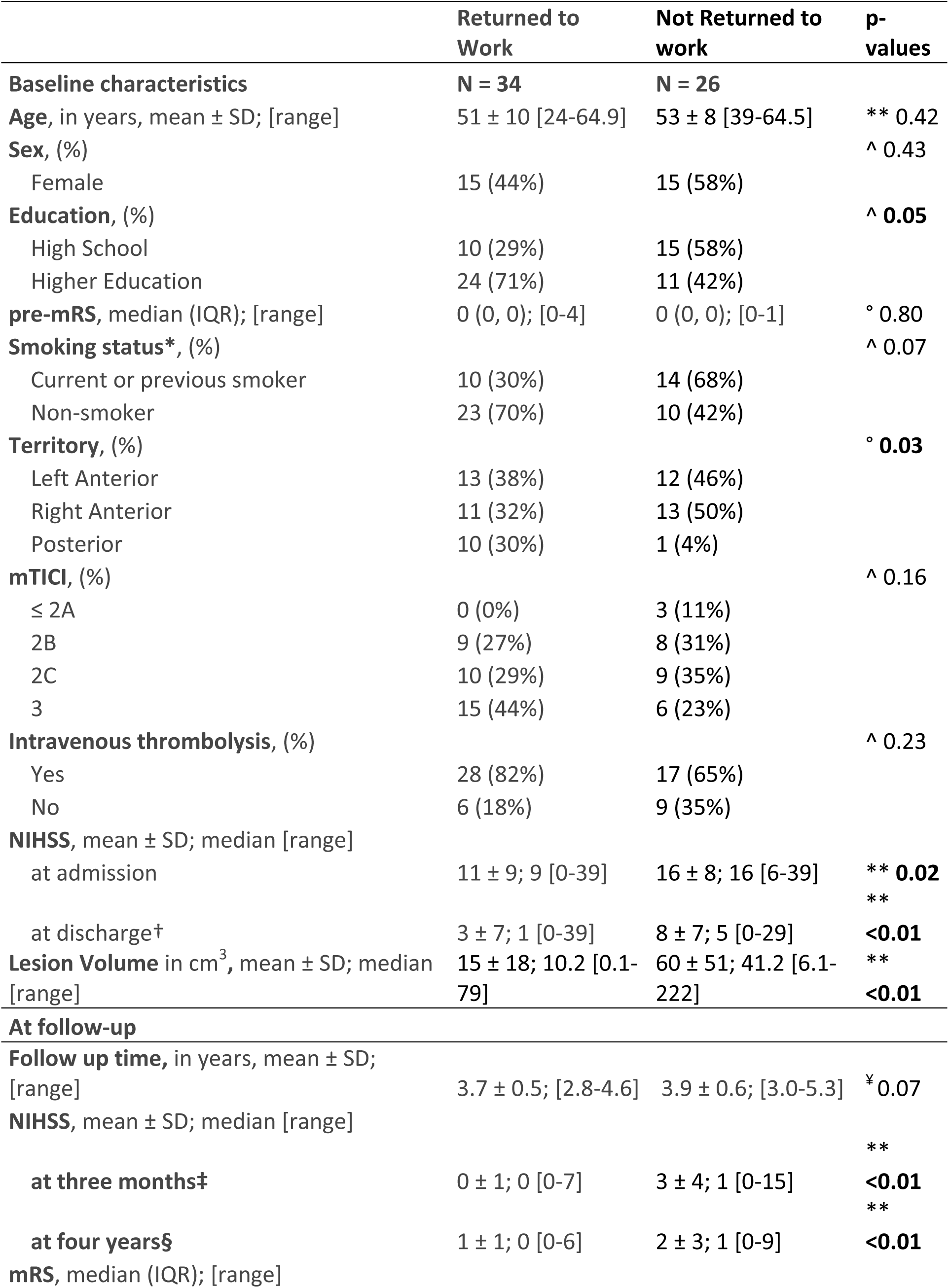
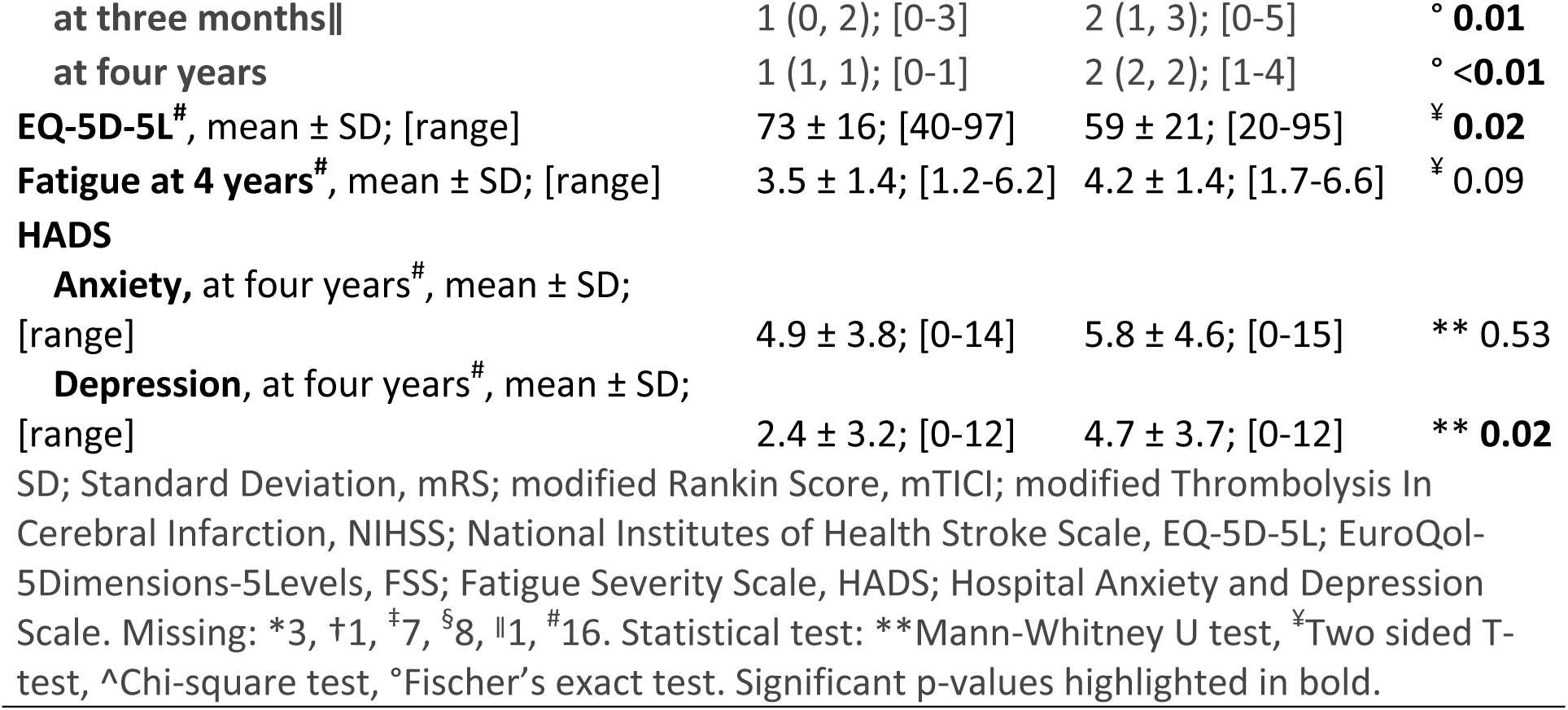
Descriptive characteristics at baseline and follow-up. N = 60.

### Prognostic accuracy of Lesion volume on NRTW

Results from the univariable and multivariable regression analyses are presented in table 2. The unadjusted estimates found a substantial and significant increase in the odds of NRTW with higher NIHSS at admission (p=0.04) and discharge (p=0.04), as well as higher lesion volume, both as a continuous (p<0.01) value and when entered as a categorical variable (p<0.01). Higher education (p=0.03) and posterior infarcts (p=0.05) were found as protective factors against NRTW.

**Table 2:** Logistic regression, association between baseline variables and RTW. Lesion volume and NIHSS was treated as continuous variables in the multivariable model. N = 60.

|  | UNIVARIABLE |  | MULTIVARIABLE |  |
| --- | --- | --- | --- | --- |
|  | Odds ratio (CI 95%) | P-value | Odds ratio (CI 95%) | P-value |
| Female sex | 1.73 (0.62 - 4.94) | 0.30 | 4.22 (0.78 - 30.1) | 0.12 |
| Age | 1.03 (0.98 - 1.10) | 0.29 | 1.07 (0.95 - 1.22) | 0.30 |
| Education High School | ref. |  | ref. |  |
| Education Higher | 0.31 (0.10 - 0.88) | **0.03** | 0.29 (0.05 - 1.35) | 0.13 |
| Vascular Territory - Left MCA | ref. |  | ref. |  |
| Vascular Territory - Right MCA | 1.28 (0.42 - 3.99) | 0.67 | 0.81 (0.13 - 4.44) | 0.81 |
| Vascular territory - Posterior | 0.11 (0.01 - 0.70) | **0.05** | 0.01 (0.00 - 0.68) | 0.10 |
| Thrombolysis | 0.40 (0.12 - 1.32) | 0.14 | 0.70 (0.08 - 5.44) | 0.73 |
| mTICI | 2.63 (0.87 - 8.70) | 0.10 | 3.99 (0.67 - 33.8) | 0.15 |
| NIHSS at admission | 1.07 (1.01 - 1.16) | **0.04** |  |  |
| NIHSS at discharge - continuous | 1.10 (1.02 - 1.23) | **0.04** | 1.11 (0.94-1.37) | 0.27 |
| NIHSS at discharge - minor | ref. |  |  |  |
| NIHSS at discharge - moderate | 7.02 (2.10 - 26.9) | **< 0.01** |  |  |
| NIHSS at discharge - severe | 8.10 (0.92 - 174) | 0.08 |  |  |
| Lesion Volume (cm<sup>3</sup>) - continuous | 1.05 (1.02 - 1.09) | **< 0.01** | 1.05 (1.02 - 1.11) | **0.02** |
| Lesion Volume (cm<sup>3</sup>) - small | ref. |  |  |  |
| Lesion Volume (cm<sup>3</sup>) - medium | 6.90 (1.90 - 29.9) | **< 0.01** |  |  |
| Lesion Volume (cm<sup>3</sup>) - large | 57.5 (8.03 - 1222) | **< 0.01** |  |  |

MCA; Middle cerebral Artery, mTICI; modified Thrombolysis In Cerebral Infarction, NIHSS; National Institute of Health Stroke Scale. Significant p-values highlighted in bold. Lesion volumes small, medium and large (<15 cm<sup>3</sup>, 15-70 cm<sup>3</sup> and >70 cm<sup>3</sup>). NIHSS at discharge minor (0-4), moderate (5-15) and severe (16-42).

After adjusting the model for age, sex, stroke territory, education, IVT, mTICI and NIHSS at discharge, lesion volume remained significant (OR 1.05, 95%CI 1.02 to 1.11, p= 0.02). No significant interaction was observed between NIHSS at discharge and lesion volume (p≍0.9). Furthermore, adjusting for intracranial volume did not alter the results for lesion volume. The results are shown in Table S5.

### Establishing a prognostic cut-off value for Lesion volume

In the univariable model, lesion volume alone showed an AUC-ROC of 0.86. The optimal cut-off for lesion volume was 29 cm^3^, yielding a sensitivity of 0.91 and a specificity of 0.65 for RTW. The positive predictive value for RTW was 78% and the negative predictive value was 85%.

### Machine learning model

The logistic regression machine learning model, as illustrated in Figure 4, yielded a mean AUC of 0.86 (SD = 0.13), demonstrating an accuracy of 75%. The model achieved a sensitivity of 0.77 and specificity of 0.72 for identifying subjects who RTW and NRTW, respectively. The permutation test saw none of its 100 iterations yield AUCs above 0.86, instead resulting in a mean AUC of 0.53 (SD = 0.10). This substantial difference in AUC’s highly suggests that the model’s predictive ability significantly outperforms random models.

**Figure 4.**
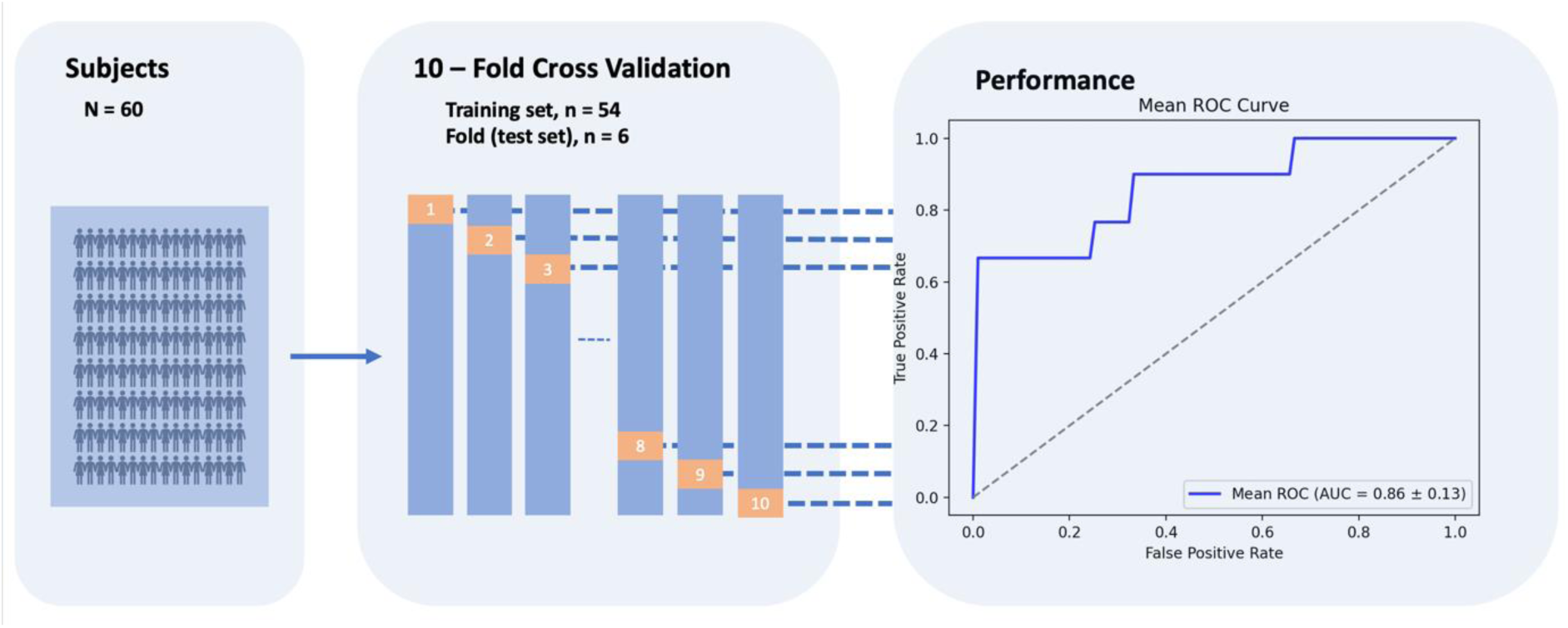
This figure provides a representation of the ten-fold cross-validation process used to validate the predictive performance of the machine learning model on not returning to work. It delineates the division into ten unique folds, each consisting of 6 subjects serving as the test set in one iteration of the validation process. The performance of the model is quantified through area under the curve, calculated as mean of folds.

Among the features considered in the model, lesion volume (mean OR=4.98) measured as a continuous variable, emerged as the most influential factor in predicting the outcome. For a comprehensive overview, we have presented the top eight most influential factors in Table S6.

## Discussion

In this study of working-age adults undergoing EVT for acute ischemic stroke, including machine learning, we found lesion volume as an imaging biomarker to be the most robust predictor of RTW. Our findings suggest that a cut-off of 29 cm^3^ is suitable to distinguish between those with high and low chance of RTW. To our knowledge, the importance of lesion volume and the use of AI to assess clinical variables on RTW have not been previously explored.

The relationship between lesion volume and RTW underscores the critical impact of stroke severity on vocational outcomes and highlights its role as a promising biomarker of RTW. Furthermore, our analysis of ML modelling demonstrated the potential to anticipate RTW outcomes based on lesion volume and other baseline variables, although this warrants further validation. The proportion of patients in this study who RTW is consistent with findings from other stroke studies.^6–8,10,33,34^ Education emerged as a protective factor for RTW. However, its significance diminished when considered alongside other variables. Contrary to previous studies, we identified an association between lesion location and RTW,^35,36^ although significance was not retained in the multivariable model. Nonetheless, a more precise exploration of localization could be valuable, as previously demonstrated by the concept of strategic locations.^37^

Previous research have uncovered the imprecise predictions of clinicians and randomness in the selection of individuals for rehabilitation.^38–40^ ML models offer a potential pathway towards a more objective, equitable and personalized stroke follow-up approach, hence reducing the undue influence of individual clinician biases. As a contribution to the growing field of stroke prediction models, we have made our pre-trained model available for download online. An obstacle to utilizing our study’s result in clinical practice is the absence of routine measurement of lesion volume. Looking ahead, the integration of AI-powered tools for image analysis holds promise for enhancing lesion volume assessments, also when it comes to regular clinical scans, such as those used in our study.^41^

Strengths of our study were the well described cohort with extensive clinical and radiological measures both at baseline and four-year follow-up. The main limitation of this study is the relatively modest sample size. Although the inclusion of diverse patients enhances generalizability, particularly to young stroke patients undergoing EVT, caution is necessary when extending our findings to broader ischemic stroke populations. Furthermore, the applicability of our results to countries with different social security systems and rehabilitation facilities than Norway should be approached with care. Additionally, several important variables were not addressed in our study, such as motivation, job type, income, rehabilitation, and the perception of social support, all of which have been established as influential factors in RTW outcomes.^8,42^ Future research could potentially benefit from obtaining the perspective of employers, as their role in facilitating RTW is a reasonable hypothesis and deserves investigation. As with other stroke prognostic machine learning models so far, another limitation is the lack of a separate test set. This diminishes certainty about the model’s generalizability, posing challenges in comparing different models and predicting their real-world performance, and is one of the reasons for making our pre-trained model available to others.

In conclusion, our study underscores the role of lesion volume as a predictor of RTW after stroke. To our knowledge, this is the first objective imaging biomarker found to be associated with RTW.

## Acknowledgements

We express our gratitude to all the patients included in this study for their time, effort, and contribution.

ChatGPT was used for improving the quality of the written English to ensure readability of the article and participated in writing the scripts for the machine learning model.

## Supplemental material list

Supplemental methods

Tables S1-S6

Reference #43 (Leonardsen EH, Peng H, Kaufmann T, Agartz I, Andreassen OA, Celius EG, Espeseth T, Harbo HF, Hogestol EA, Lange AM, et al. Deep neural networks learn general and clinically relevant representations of the ageing brain. *Neuroimage*. 2022;256:119210. doi: 10.1016/j.neuroimage.2022.119210)

## Non-standard abbreviations and acronyms

AI: Artificial Intelligence
DWI: Diffusion Weighted Imaging
EVT: Endovascular treatment
IVT: Intravenous Thrombolysis
MCA: Middle Cerebral Artery
ML: Machine Learning
MRI: Magnetic Resonance Imaging
mRS: Modified Rankin Scale
NIHSS: National Institute of Health Stroke Scale
NRTW: Not Return to Work
OR: Odds Ratio
OSCAR: The Oslo Acute Reperfusion Stroke Study
RTW: Return to Work
mTICI: modified Thrombolysis in Cerebral Ischemia

